# Normal mean oral temperature is 98°F, not 98.2°F or 98.6°F

**DOI:** 10.1101/2021.06.03.21258282

**Authors:** Nitin Kumar, Kavya Ronanki, Prasan Kumar Panda, Mayank Kapoor, Yogesh Singh, Ajeet Singh Bhadoriya

## Abstract

**Background:** 98.6°F is generally accepted as normal body temperature as defined by Wunderlich (1868) and later challenged by Mackowiak (1992) and Protsiv (2020) who concluded as 98.2°F based on cross-sectional studies. Hence, the normal body temperature at present needs quantification?

**Methods:** A longitudinal study on the healthy population of Northen-India were followed-up over 1-year. Participants were advised for self-monitoring of oral temperature with a standard digital thermometer in either left or right sublingual pocket and record it in the thermometry diary. The study was considered complete if the participant had all the three phases of the study (i.e. non-febrile, febrile, and post-febrile phases) or completed the duration of the study.

**Results:** The mean oral temperature of the participants (n=144) during the non-febrile and post-febrile phases (temperature readings=6543) were 98°F (SD, 0.61) and 98.01°F (SD, 0.60) respectively (P<0.001). The mean oral temperature during post-febrile phase was found to be 0.01°F higher than non-febrile phase. With the diurnal variability, the morning (AM), noon (AN), and afternoon (PM) mean temperatures were 97.91, 98.08, and 98.27°F (P<0.001) respectively during the non-febrile phase. Similar trends were observed in variability among men and women, and seasons.

**Conclusions:** The mean oral temperature is 98°F (SD, 0.61). The temperature is as low as 96.9^0^F and as high as 99.1^0^F. The temperature during post-febrile phase was found to be higher than the non-febrile phase temperature like PM over AN & AM, women over men, summer over other seasons in the non-febrile phase, spring over others in the post-febrile phase.

## Introduction

Maintaining normal body temperature is vital to health, the measurement of which is critical while evaluating a patient as it can indicate a significant life-threatening illness. Although the entrenched meaning of fever is an increase in body temperature, the exact scientific definition is still debatable. It can never be defined without exactly defining what is the normal temperature in the human body. Wunderlich (1868) defined the normal body temperature as 37°C (98.6°F). He measured temperature in the axilla of patients, i.e. non-healthy participants in a cross-sectional study, but the axillary temperature is far representative of core body temperature (1)(2). Furthermore, the thermometer Wunderlich used was not standardized and might have resulted in higher values compared with modern digital thermometers. He defined normal body temperature when life expectancy was low (i.e. 38 years) and the prevalence of untreated chronic infections such as tuberculosis, syphilis, and periodontitis was high (3–5). It was an over-generalization to a healthy population (1). His results might have been influenced by several confounders, e.g. age, sex, race, menstruation, pregnancy, day-night (diurnal), seasons, climates, and altitudes.

Since modern temperature guidelines are based on age-old data, Mackowiak, Wasserman, and Levine (1992) set out to question this time-honored Wunderlich’s dictum. They did their cross-sectional study on young adults (younger than 40 years) using a standardized thermometer (1). They concluded that 36.8°C (98.2°F) rather than 37.0°C (98.6°F) was the mean oral temperature of their participants; 37.7°C (99.9°F) rather than 38.0°C (100.4°F) was the upper limit of the normal temperature range. However, the most vulnerable younger and older populations were not included in the study. Protsiv et al. (2020) hypothesized that the normal oral temperature of adults is lower than the established 37°C of the 19th century and concluded that body temperature has decreased over time in the USA using measurements. Recent studies suggest that normal temperature has invariably decreased by 0.03°C per birth decade probably due to lowered metabolic rate and infections, henceforth drifting down the normal morning body temperature to less than <98.6°F over the last two centuries (6–10). The influence of age, time of day, gender, and economic development preclude an updated definition of normothermia.

Most importantly, all studies till now were cross-sectional resulting in a complete bias of the measured temperature whether in pre-febrile, febrile, or post-febrile phase. And being raised temperature above normal (known as fever) is a sign that should have been studied longitudinally. We hypothesized that the core body temperature has decreased which is a crude surrogate for basal metabolic rate. The change over time provides important physiologic clues to alterations in human health.

Considering 98.6°F as normal body temperature in the light of newly available evidence would have untoward consequences, and it has been riddling since the inception of modern medicine and needs to be relooked into a new dimension preferably through a developing society. This can only be answered via a prospective study with monitoring of body temperature on a healthy population when they are afebrile, on follow-up attain fever, and on their post-febrile phase.

Thereby, we did a longitudinal study on healthy participants from a tertiary care institute, using a standardized electronic thermometer in the left or right posterior sublingual pocket to determine the mean oral temperature during the non-febrile phase and to determine the diurnal, gender, age-related, and seasonal variations of oral temperature in the same population during non-febrile, febrile, and post-febrile phases on follow-up (2,11).

## Methods

### Study setting, design, and participants

The study was conducted at All India Institute of Medical Sciences Rishikesh (AIIMS), a tertiary healthcare center in the state of Uttarakhand, India. It was a longitudinal study conducted over 15 months from July 2019 to September 2020.

Lists of employees and students were obtained from the human resource department and registrar’s office. Information of participating family members was obtained from consenting employees and students and was selected via a simple random sampling method using the computer. If the participant did not give consent for the study then the next person on the list was selected. We had taken the standard deviation according to a study done by Mackowiak et al as 0.7 and employing T-distribution to estimate sample size, the study would require a sample size of 192 with 95% confidence and a precision of 0.1 (1,12). By adjusting non-responders of 20 %, the final sample size was 232. Due to the ongoing COVID-19 pandemic, it was restricted to 215.

The participants were recruited based on the following inclusion and exclusion criteria after taking informed consent.

- Inclusion criteria: healthy staff and students of AIIMS and their accompanying family members between 4-100 years and who agreed to work/study at AIIMS during the duration of the study (1year).
- Exclusion criteria: any individuals with any diagnosed or suspected disease (any acute infectious or non-infectious illness (including trauma) within last 1month and post-partum period up to 8 weeks; any known case of or past history of chronic illness - infective (e.g. tuberculosis, kala-azar, brucellosis, infective endocarditis, HIV/AIDS, hepatitis B/C/D etc.), rheumatological (e.g. RA, SLE, vasculitis etc.), chronic liver disease, chronic kidney disease, cardiovascular disease (e.g. systemic hypertension, coronary artery disease, valvular heart disease, pulmonary arterial hypertension, peripheral vascular disease etc.), chronic lung disease (e.g. any obstructive or restrictive airway diseases), endocrinopathy (e.g. diabetes mellitus, diabetes insipidus, hypo/hyperthyroidism etc.), gastro-intestinal disease (e.g. dyspepsia, inflammatory bowel disease, malabsorption syndromes etc.), neurologic disorders (e.g. epilepsy, stroke, dementia, movement disorder, degenerative disorder, cerebral palsy etc.), psychiatric disorder (e.g. mood disorders, psychosis, dependence syndrome(s) etc.), dermatological diseases (e.g. bullous disorders, psoriasis, tinea etc.), any malignancy (treated or otherwise), recent history of vaccination in last 6 months, and ankyloglossia.

### Interventions

Detailed clinical evaluations (history and examination) were done. Basic investigations (with the last 1year; as per institute recruitment policy): ECG, chest X-ray, viral markers (anti-HIV-1 and 2, HBsAg, anti-HCV), urine routine, complete blood count, fasting blood glucose, liver and kidney function tests were collected from medical record section after approval from Institute Ethical Committee (No. 235/IEC/PGM/2019). If any abnormality was detected, they were excluded without sharing details (Fig 1).

**Fig. 1:**
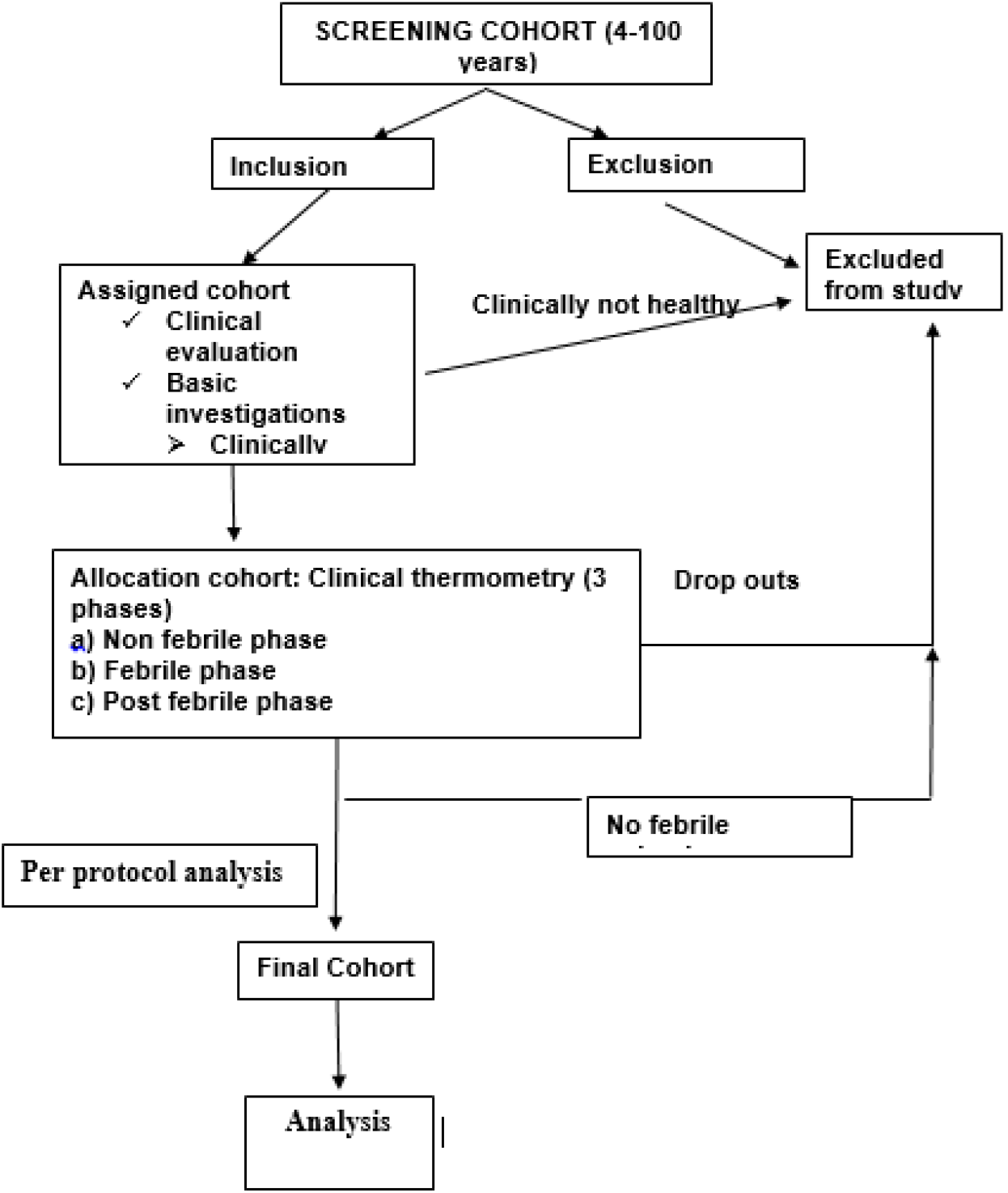
The study flow.

The study was done in three phases: first phase (non-febrile phase), second phase (febrile phase), and third phase (post-febrile phase). One clinical thermometry diary, a ball pen, a and standard electronic thermometer (Dr.Morepen Digiflexi Flexi Tip Thermometer (MT222)) were provided to all participants. The detailed procedure was explained the and first reading was verified and they were followed up fortnightly physically, and reminded weekly through WhatsApp and telephonically. Procedure to be followed while measuring temperature was explained as follows:

- Wash your hands with soap and water.
- Use a clean thermometer, one that has been washed in cold water, cleaned with rubbing alcohol, and then rinsed to remove the alcohol.
- Please ensure that no physical exertion in the past 30min, avoid any cold or hot beverage or food in the past 30min, no smoking during the past 30 min.
- Sit in a cool and calm environment for at least 30 min.
- Keep the thermometer in an oral left or right posterior sublingual pocket and keep your mouth closed during this time.
- Hold the thermometer in the same spot until it makes a beeping noise when the final reading is done.
- Readings will continue to increase and the °F (& °C) symbol will flash during measurement.
- Kindly record the temperature on the Fahrenheit scale and the time in the thermometry diary.
- Rinse the thermometer in cold water, clean it with alcohol, and rinse again and store it in a safe place for the next readings.
- Three readings were taken once after waking up (AM), once in the afternoon (AN; 12-3 PM), and once before sleeping (PM), also requested to mark any symptoms from the checklist simultaneously.
- There were 3 days of more frequent temperature charting (every 2^nd^ hour, except sleeping time) during the non-febrile phase and 2 days of frequent temperature charting during post febrile phase, and for all the days during the febrile phase
- For family members having children and old age persons, staff or students measured the temperature and recorded.

Self-recording of data was done in the provided clinical thermometry diary and the same was assessed fortnightly by the investigator(s) for troubleshooting and to see the status of recording. Diary was collected once the participant had all the 3 phases of the study or at the end of the study if no febrile episodes.

### Comparator/outcomes

There was no comparator except among three phases of temperature recordings. The data was entered in the excel sheet and primary outcomes were analyzed as per-protocol analysis (for those participants who had all the three phases in the study) using Statistical Package for Social Sciences (SPSS) Version 23.

Participants with the febrile phase were further divided into 4 subgroups based on seasonal months: November-January (represented coldest months of the year); February-April (representing spring months); May-July (representing hottest months); August-October (representing autumn months). A maximum of 45 days of data was taken immediately before the febrile phase, during the non-febrile phase of per-protocol analysis. A maximum of 10 days of data was taken immediately after the febrile phase during the post-febrile phase, and complete data of the febrile phase was taken for analysis. The whole of the frequent temperature readings (i.e. 2 hourly temperature records) was taken for analysis for all the 3 phases especially to see variations.

### Statistical analysis

Categorical variables were presented in number and percentage (%) and continuous variables were presented as mean (with SD) and median (with IQR). Normality of data was tested by Kolmogorov-Smirnov test. If the normality was rejected then a non-parametric test was used. Statistical tests were applied as follows:

i. Quantitative variables were compared using the Independent t-test/Wilcoxon–Mann– Whitney test (when the data sets were not normally distributed) between the two groups and the Kruskal-Wallis test between three and more groups.
ii. Qualitative variables were correlated using the Chi-Square test/Fisher’s Exact test.
iii. Relationships were assessed using Pearson or Spearman tests depending upon distribution.
iv. The continuous variables, that were not normally distributed were analyzed using Shapiro-Wilk Test.

Taking confidence level as 95%, the statistical significance was defined as P <0.05 significant.

## Results

350 participants were screened, 250 consented to be a part of the study, and 215 found to be clinically healthy. Nineteen of the participants did not complete the study, 52 had no febrile phase during the study period, thus excluded from the outcome analysis. 144 had a febrile phase ranging from 2 to 8 days and a post-febrile phase. 52.1% of participants were male. The majority of subjects were from 10 to 40 years (97.64%). Student (MBBS and nursing) constituted 58.1%, doctors 32.1%, rest were other staffs. Demographic profile, as well as some important physiological variables, were described in table 1.

**Table 1:** Physiologic and biochemical characteristics of the participants at first contact (n=215)

|  |  |  |  |
| --- | --- | --- | --- |
| Parameter | Mean ± SD | Median (IQR) | Min - Max |
| Age (Years) | 24.24 ± 5.92 | 23.00 (20.00-27.00) | 8.0 - 58.0 |
| Systolic BP (mmHg) | 114.91 ± 8.77 | 116.00 (110.00-121.00) | 96.0 - 132.0 |
| Diastolic BP (mmHg) | 73.72 ± 8.78 | 72.00 (68.00-80.00) | 58.0 - 90.0 |
| Pulse Rate (bpm) | 79.09 ± 9.18 | 78.00 (72.00-89.00) | 57.0 - 104.0 |
| Respiratory Rate (cpm) | 16.76 ± 2.28 | 17.00 (15.00-18.00) | 12.0 - 22.0 |
| Temperature ($^{\circ}$F) | 98.07 ± 0.56 | 98.0 (97.90-98.60) | 96.9 - 99.1 |
| **Hemoglobin (gm/dL)** | 14.10 ± 1.15 | 13.90 (13.15-15.00) | 12.5 - 16.5 |
| **TLC (/cu.mm)** | 7508.55 ± 2043.22 | 7404.00 (5734.00-9427.50) | 4024.0 - 10988.0 |
| **Platelet Count (lakh/cu.mm)** | 2.54 ± 0.63 | 2.60 (2.00-3.10) | 1.5 - 3.5 |
| **FBS (mg/dL)** | 85.85 ± 9.31 | 85.00 (79.00-94.00) | 70.0 - 101.0 |
| **T. Bilirubin (mg/dL)** | 0.69 ± 0.32 | 0.70 (0.40-1.00) | 0.2 - 1.2 |
| **D. Bilirubin (mg/dL)** | 0.19 ± 0.08 | 0.20 (0.10-0.30) | 0.1 - 0.3 |
| **SGPT (IU/L)** | 31.43 ± 14.05 | 32.00 (20.00-43.00) | 7.0 - 56.0 |
| **SGOT (IU/L)** | 25.13 ± 11.96 | 27.00 (14.50-35.00) | 5.0 - 46.0 |
| **S. Albumin (gm/dL)** | 4.26 ± 0.45 | 4.30 (3.90-4.60) | 3.5 - 5.0 |
| **S. Urea (mg/dL)** | 24.30 ± 8.93 | 24.00 (17.00-31.00) | 10.0 - 40.0 |
| **S. Creatinine (mg/dL)** | 0.69 ± 0.31 | 0.70 (0.40-1.00) | 0.2 - 1.2 |
| **S. Sodium (mEq/L)** | 139.89 ± 2.99 | 140.00 (137.00-142.00) | 135.0 - 145.0 |
| **S. Potassium (mEq/L)** | 3.97 ± 0.34 | 4.00 (3.70-4.30) | 3.4 - 4.5 |

### Per-protocol analysis (n=144)

The normal temperature variation was measured (Table 2)

**Table 2:**
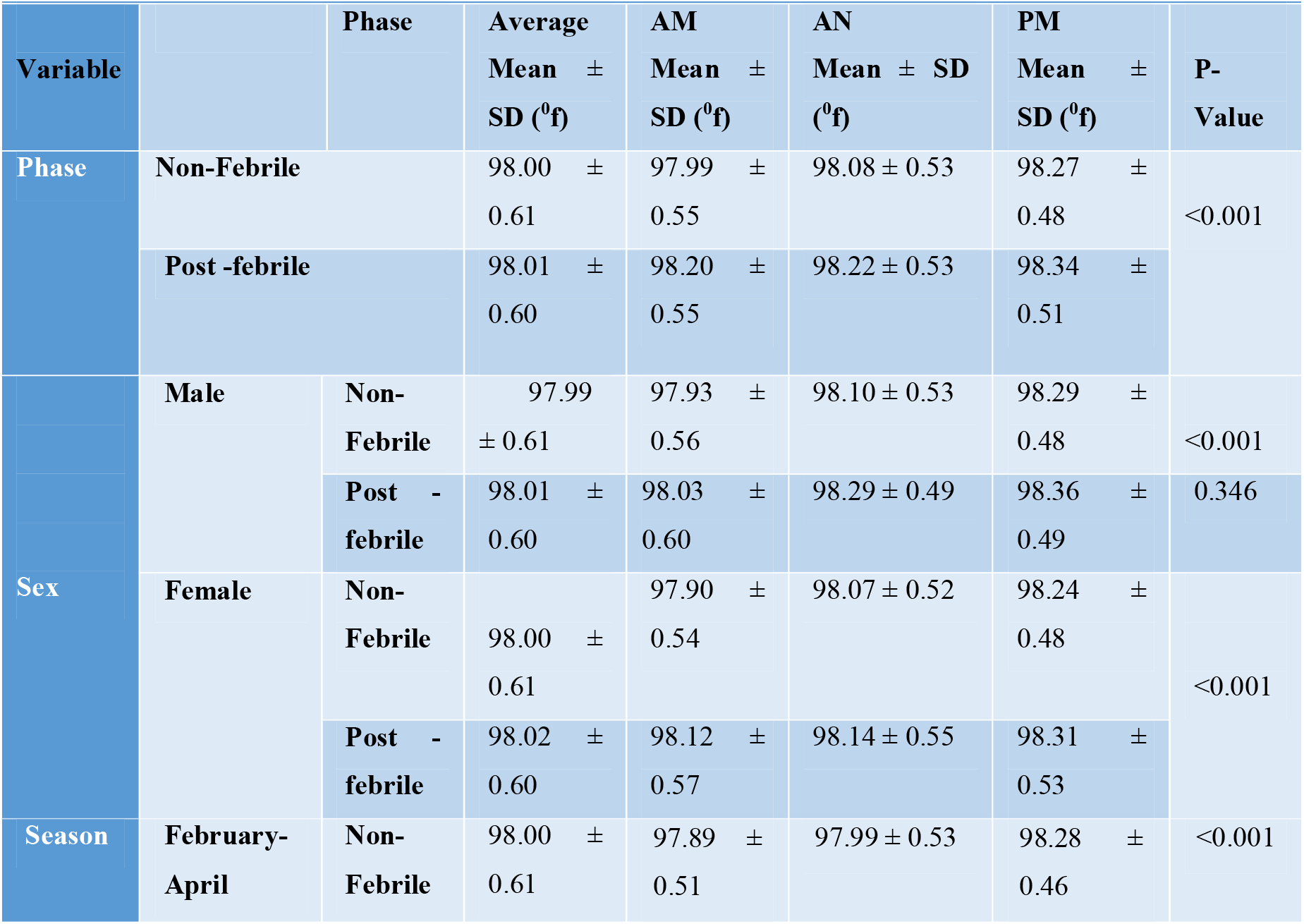
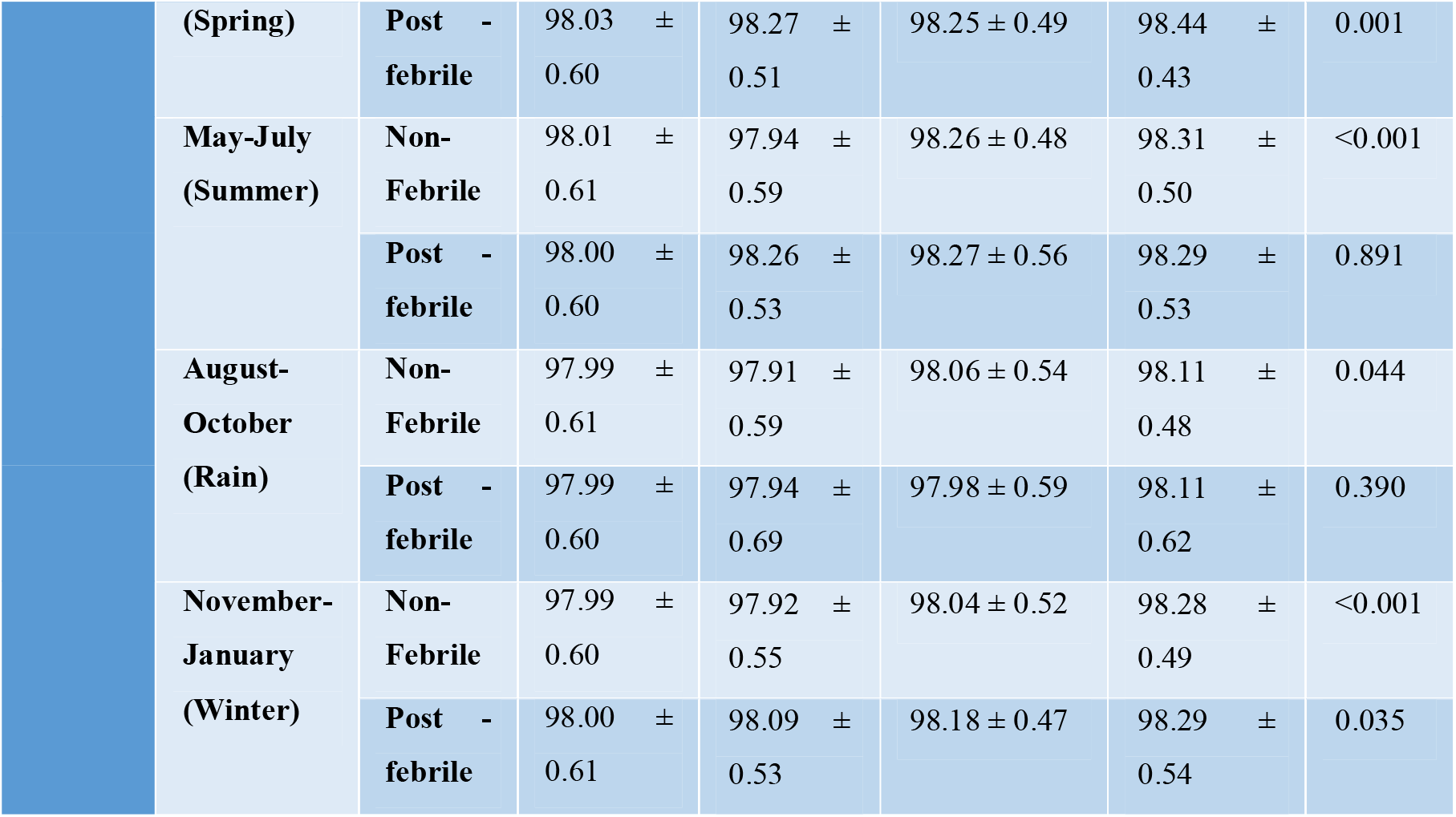
Normal variation of temperature in a healthy cohort.

## Discussion

We analyzed 6544 temperature readings of the 144 healthy participants longitudinally over 1-year. The mean oral temperature was 98^0^F (SD 0.61) with a range of 96.9 – 99.1^0^F. However, 98.6^0^F (SD, 0.36) is generally considered physiologic convention (13). The variations in oral temperature in our study are far exceeding the physiologic convention which was generated from analyzing the western population to define physiologically accepted convention and by marginalizing geographic, local, racial, and epidemiological factors. It can be considered that the normal range of temperature is more varied in the tropics, and the normal body temperature can be considered high in a tropical climate and so the variation. The higher temperature found in our study can be attributable to climate and relatively high incidence and prevalence of acute and chronic infections respectively. Western countries have eliminated (controlled) many acute and chronic infectious diseases, and infectious diseases are one of the determinants of body temperature (5). The above normal temperature found in our study agrees with the study done by Renbourn and Bonsall (1946) (14).

According to the present study, the temperature as low as 96.9^0^F and as high as 99.1^0^F can be considered normal to a significant proportion of the populace. However, presently we are following 99.9°F is the upper limit of normal temperature that was previously was 100.4°F (1). This upper limit is too lowered in the present study, leads a significant development in the history of medicine. This change reflects what Protsiv et al concluded that men born in the early 19^th^ century had temperatures 0.59°C higher than men of the 20^th^ century, with an invariable decrease of 0.03°C per birth decade (6). So does the temperature has decreased in women by 0.32°C during the same time frame with a similar rate of decline (0.029°C per birth decade). It can be attributed to decreasing incidence and prevalence of both acute and chronic infections respectively, improved with the dawn of antibiotic age together, improved standards of living, living in the thermo-neutral environment.

The mean oral temperature during the post-febrile phase was found to be 0.01^0^F higher than the non-febrile phase temperature (98.0^0^F; SD, 0.61). It is difficult to delineate the time required to return pre-febrile temperature, as it will require a larger sample size and longer follow-up to say with certainty. However, the same observation was observed in past too. Regarding seasonal variation, the mean oral temperature was found to be highest during the summer season (98.01^0^F; SD, 0.61), it would not be wrong to consider that the environmental temperature does affect the body temperature. However, This couldn’t hold for post-febrile phase physiology as the maximum mean temperature was found to be highest (98.03^0^F; SD, 0.60) during the spring season (pre-summer). The variation can be attributable to maximum readings owing to the maximum incidence of fever during the spring season. No previous studies have observed these changes.

The mean oral temperature was found to be higher in women compared to men during the non-febrile and post-febrile phases. This represents physiological raised temperature in ovulating cycles in women that was not studied in the present study. The AM-AN-PM variability in the mean of oral temperature was in concordance with accepted physiological convention i.e. the body temperature arises as day passes (15).

Being a longitudinal study, our study overcomes some of the drawbacks of previous studies; it has got some limitations too. The sample was unicentric, difficult for generalization of results, thus a larger multi-centric study is required. The vulnerable group of the population, elderly and children, could not be included in the study desirably. The participants were defined healthy based on history and pre-defined biochemical and laboratory parameters, therefore, indolent chronic infections and sub-clinical non-infectious illnesses couldn’t be ruled out. No physical way of checking the adherence to the advised procedure for the temperature measurement was there. The participants were reviewed followed up fortnightly. So, strategies to measure directly observed temperature may require. Compliance was overall poor (∼38%). The oral temperature in the left or right sub-lingual pocket is close representative of core body temperature, but not exactly the core body temperature. The ongoing COVID-19 pandemic might have influenced the results. The participants constituted a high-risk populace for infectious agents and stress. The etiology of febrile illnesses cannot be solely attributable to infectious agents. The etiologies have to be looked at in future studies to determine the strength of infectious agents in the regulation of core body temperature. The time frame for the restoration of normal physiology cannot be ascertained. We didn’t record the exact ambient indoor temperature.

In conclusion, this study revealed that the mean oral temperature (during the non-febrile phase) is 98^0^F, not 98.6^0^F. The temperature is as low as 96.9^0^F and as high as 99.1^0^F, which can be considered normal to a significant proportion of the populace. The temperature during the post-febrile phase was found to be higher than the non-febrile phase temperature like women over men, summer over other seasons in the non-febrile phase, spring over others in the post-febrile phase. The AM-AN-PM variability of temperatures was in concordance with accepted physiological convention but did not hold for the febrile phase. Knowledge regarding the accurate normal values is needed; hence, further multicentric international study is needed to define the normal range of this vital sign, the temperature.

## Data Availability

It is available with the corresponding author, once required, will be provided.

## Contributors

NK contributed to the data collection, data analysis, and was involved in manuscript writing. KR, MK, YS, and ASB contributed to the data analysis and were involved in reviewing the draft. PKP gave the concept, interpreted analysis, critically reviewed the draft, and approved it for publication along with all authors.

## Data sharing

It will be made available to others as required upon requesting the corresponding author.

## Acknowledgment

None

